# SETBP1 Haploinsufficiency and Related Disorders Clinical and Neurobehavioral Phenotype Study

**DOI:** 10.1101/2024.05.23.24307620

**Authors:** Haley O. Oyler, Caitlin M. Hudac, Wendy K. Chung, LeeAnne G. Snyder, Stephanie Robertson, Siddharth Srivastava, Trina Geye

**Affiliations:** SETBP1 Society, Austin, TX 78703; Department of Psychology and Carolina Autism and Neurodevelopmental Research Center, University of South Carolina, Columbia, SC, 29201; Department of Pediatrics, Boston Children’s Hospital and Harvard Medical School, Boston, MA 02115; Simons Foundation, New York, NY, 10010; Associate Professor of Psychological Sciences and Director of the Tarleton Center for Child Well-being, Tarleton State University, Stephenville, TX 76402; Department of Neurology, Boston Children’s Hospital and Harvard Medical School, Boston, MA 02115; Associate Professor of Psychological Sciences, Tarleton State University, Stephenville, TX 76402

**Author notes:** Corresponding Author: Haley Oyler.

## Abstract

To comprehensively investigate the neurodevelopmental profile and clinical characteristics associated with SETBP1 haploinsufficiency disorder (SETBP1-HD) and SETBP1-related disorders (SETBP1-RD). We reported genetic results on 34 individuals, with behavior and clinical data from 22 with SETBP1-HD and 5 with SETBP1-RD, by assessing results from medical history interviews and standardized adaptive, clinical, and social measures provided from Simons Searchlight. All individuals with SETBP1-HD and SETBP1-RD exhibited neurological impairments including intellectual disability/developmental delay (IDD), attention-deficit/hyperactivity disorder, autism spectrum disorder, and/or seizures, as well as speech and language delays. While restricted interests and repetitive behaviors present challenges, a relative strength was observed in social motivation within both cohorts. Individuals with SETBP1-RD reported a risk for heart issues and compared to SETBP1-HD greater risks for orthopedic and somatic issues with greater difficulty in bowel control. Higher rates for neonatal feeding difficulties and febrile seizures were reported for individuals with SETBP1-HD. Additional prominent characteristics included sleep, vision, and gastrointestinal issues, hypotonia, and high pain tolerance. This characterization of phenotypic overlap (IDD, speech challenges, autistic and attention deficit traits) and differentiation (somatic and heart issue risks for SETBP1-RD) between the distinct neurodevelopmental disorders SETBP1-HD and SETBP1- RD is critical for medical management and diagnosis.

## INTRODUCTION

### SETBP1

*SETBP1* (HGNC:15573)*, SET binding protein 1* is a dosage-sensitive gene located at 18q12.3 that encodes a regulatory protein essential to human brain development.^1,2^ SETBP1 functions as a transcription factor involved in cell cycle control, gene expression through modulation of chromatin accessibility, and regulation of transcription and is implicated in speech and language development.^2,3,4^

### SETBP1-related Neurodevelopmental Disorders

Variants in *SETBP1* are associated with three different phenotypes: 1. Schinzel-Giedion Syndrome (SGS), 2. SETBP1 haploinsufficiency disorder (SETBP1-HD), and 3. SETBP1- related disorders (SETBP1-RD).

SGS (OMIM #269150) is a rare, often progressive, neurodevelopmental disorder characterized by distinct facial features, skeletal, neurological, cardiovascular, and congenital anomalies with increased risk of malignancy.^5,6,7^ SETBP1 variants associated with SGS are isolated to a specific amino acid region on SETBP1 (868-871).^8^ Prior literature identified a sub-category called atypical SGS for individuals with variants located adjacent to or close by the classical SGS amino acid region (862-867,872-873) with less severe symptoms and a potentially longer life expectancy, past the first decade in life.^6^ Diagnosis is often made initially based on physical, clinical presentation and verified with genetic testing.

SETBP1-HD (OMIM #616078) is a rare neurodevelopmental disorder characterized by speech/language impairment , intellectual disability/developmental delay (IDD/DD), hypotonia, attention deficits, vision impairment, characteristic facial features, and autistic traits.^1,4,9^ SETBP1 variants associated with SETBP1-HD span the length of the *SETBP1* gene. Although a prior publication noted some subtle and less recognizable facial differences in 93% of individuals with SETBP1-HD which are distinct from SGS, including ptosis, broad nasal bridge, hypertelorism, full nasal tip, high palate and blepharophimosis, the differences are not sufficiently distinct for a clinical presentation diagnosis and therefore diagnosis of SETBP1-HD is determined by genetic testing.^9^

In general, loss of function SETBP1 variants are associated with SETBP1-HD, whereas gain of function SETBP1 variants are associated with SGS. A third, novel category comprises variants associated with SETBP1-RD due to pathogenic missense variants located outside the classical and atypical SGS region (862-873) or in-frame deletions, with DNA-binding and transcription function independent of SETBP1 protein abundance.^6,10,11^ All SETBP1-related neurodevelopmental disorders are autosomal dominant and primarily de novo, with rare cases resulting from inherited mutations.^10,12^

### Summary

Knowledge is limited for the neurodevelopmental impact and phenotypic consequences of *SETBP1* changes, particularly for variants that lead to SETBP1-HD and SETBP1-RD.^1,2,10,13^ Here, we present the first comprehensive cross-sectional and longitudinal, when available, comparative summary of behavioral and clinical phenotypes for SETBP1-HD and SETBP1-RD. This paper includes neurodevelopmental impact and phenotypic characteristics for 22 cases;12 previously unpublished cases of SETBP1-HD and 6 previously unpublished cases of SETBP1- RD. Limited extant literature describes the neurodevelopmental profile and clinical characteristics of individuals with SETBP1-HD and SETBP1-RD, which critically impairs understanding of the disorders, thereby hindering access to an accurate diagnosis, proper care, and medical management.^4,7,10,11,14^ To date, there are no treatments specific to SETBP1-HD or SETBP1-RD; a well-established neurodevelopmental and clinical profile of the disorders is a vital step on the path to developing standard of care guidelines and targeted treatments for affected individuals.

## MATERIALS AND METHODS

### Simons Searchlight Data Collection

For this publication, we analyzed Simons Searchlight (Searchlight) data from participants with SETBP1 variants. Searchlight is an international research program by the Simons Foundation Autism Research Initiative (SFARI) aiming to characterize the natural history of rare genetic neurodevelopmental disorders caused by a single gene or copy number variant. Caregivers register online with Searchlight, then complete a medical history screener and interview (MHI) with a licensed genetic counselor as well as a series of standardized caregiver-reported questionnaires online. Genetic reports are uploaded and variants are confirmed for pathogenicity. Many of the online instruments are administered annually to obtain longitudinal data for participants. SETBP1 Society is a 501(c)3 non-profit with a mission to provide support for families affected by SETBP1-HD and SETBP1-RD. The society functions to promote and fund research, bring awareness and education to the public, and to unite the community. One way the society upholds its mission is through the promotion and recruitment for Searchlight. As an extension of its mission, the organization partnered with Tarleton State University to develop the SETBP1 Community Research Study (SCoReS) study. SCoREs is a community based participatory research (CBPR) project designed to be iterative and ongoing with a focus on intervention development with equal partnership between the community and the researcher team.^15^ The SETBP1 Searchlight v10 and v11 data for this publication were extracted from the SFARI Base database on July 7, 2022 and January 24,2023.

### Verification and Classification

Of the 34 individuals with a SETBP1-HD or SETBP1-RD diagnosis, whole exome sequencing (WES) was the most common method for identifying SETBP1-HD and SETBP1-RD, 80% of the participants. The additional genetic tests that identified the diagnosis were Chromosomal microarray analysis (CMA) and whole genome sequencing (WGS), as well as targeted panels, including the Autism/ID Xpanded Panel, EpiXpanded Panel, Epileptome and Congenital Hypotonia Xpanded Panel.

We included participants with pathogenic or likely pathogenic SETBP1 variants or deletions who were categorized into one of two groups: SETBP1-HD, comprising individuals with SETBP1 loss-of-function (nonsense, frameshift) variants or with intragenic or full deletions of SETBP1 affecting up to 2 adjacent genes, and SETBP1-RD, comprising individuals with missense SETBP1 variants located outside the classical and atypical SGS region (862-873). Individuals with a Variant of Uncertain Significance (VUS), SGS or SETBP1 groups (deletions encompassing SETBP1 and 3 or more additional genes or splice site variants) with less than 3 individuals with data were excluded. For a complete list of SETBP1 variants included in Searchlight, see Supplementary Table 1.

### Standardized Measures

The Vineland Adaptive Behaviors Scales, third edition (VABS-3) assesses adaptive functioning necessary for individuals to live as independently and to function as safely and appropriately in daily life as possible, given their age.^16^ Adaptive behaviors include practical skills like grooming, safety, following rules, ability to work, taking care of oneself and their environment, making friends, social ability, and personal responsibility. This standardized measure has motor, communication, daily living skills, and socialization domains, resulting in an overall Adaptive Behavior Composite (ABC) score.

The caregiver-reported Child Behavior Checklist (CBCL) uses a 3-point Likert scale to assess an array of behavioral and emotional concerns in children by providing broadband, narrowband, and DSM-5 oriented scales.^17,18,19^ Both the Child Behavior Checklist 1½ -5 (CBCL/1½−5) for preschool children and the Child Behavior Checklist 6-18 (CBCL/6-18) for school-age children were administered within Searchlight based on developmental level and appropriateness.

The Social Responsiveness Scale, Second Edition (SRS-2) School Age measures social deficits and other symptoms related to autism spectrum disorder (ASD) observed in natural settings, using a 4-point Likert scale.^20^ The SRS-2 provides a total score and five treatment subdomain scores: social awareness, social cognition, social communication, social motivation, and restricted interests and repetitive behaviors.,

The Social Communication Questionnaire – Lifetime (SCQ) is a caregiver-report screening measure that assesses the presence or absence of symptomatology associated with ASD for children over the age of 4 years.^21^ The SCQ yields a single total score with a cutoff score of >= 15, used as an indication of a possible ASD diagnosis and indicates the need for further comprehensive evaluation.

The Children’s Sleep Habits Questionnaire (CSHQ) is a caregiver-report tool developed to evaluate sleep patterns and problems in children aged 2–17 years using a 3-point Likert scale.^22,23^ A total score of 41 is reported to be a sensitive clinical cutoff score for identifying probable sleep problems. Eight subscale scores are calculated with higher scores for these subscales indicating more sleep problems.

Additional Searchlight surveys reviewed included the Seizure Survey, Sleep Supplement, Brief Developmental Update and Background History Form.

When sufficient longitudinal data were reported for the standardized measures, as determined by more than half of caregivers completing the survey more than once, all data points were considered for age-relevant trends with statistical controls (i.e., random intercept per person) to account for the multiple time points. If only cross-sectional data were reported for specific surveys, then the most recent evaluation was included for each individual.

## RESULTS

### Genotype & Demographics

Of the 58 participants with genetic reports associated with SETBP1 in Searchlight, 28 individuals have a confirmed diagnosis of SETBP1-HD; 16 male and 12 female. Individual 17 had an additional likely pathogenic variant in the *CHD7* gene, inherited from a parent, who did not exhibit the symptoms of CHARGE syndrome, which is a separate disorder that can be caused by pathogenic CHD7 variants. Individual 28 had a likely pathogenic variant in the *SDHB* gene but was not included in the phenotypic analysis due to missing data. No other additional pathogenic variants were determined in the remaining 26 participants in the SETBP1-HD group. Six individuals had a pathogenic or likely pathogenic missense SETBP1 variant located outside the atypical and classical SGS region and comprised the SETBP1-RD group (Table 1). In the case where the inheritance is known, all SETBP1 variants associated with SETBP1-HD and SETBP1-RD were reported as de novo.

**Table 1.**
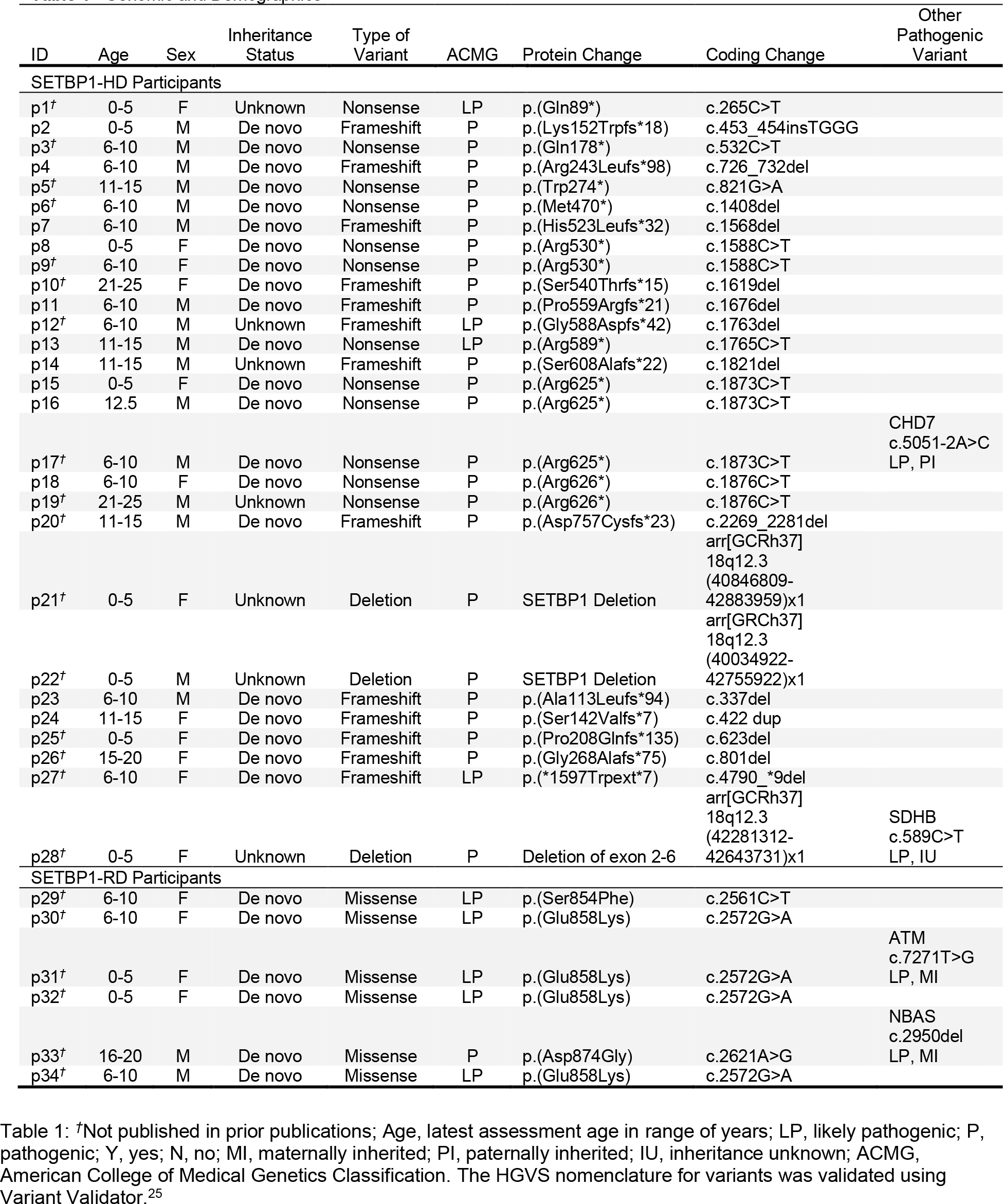
Genomic and Demographics.

Twelve of the 28 individuals with SETBP1-HD had a confirmed nonsense variant while 13 had a frameshift variant and 3 had a partial deletion of the SETBP1 gene. All of the deletions and variants were unique except for 3 individuals with the p.(Arg625*) variant, 2 individuals with the p.(Arg530*) variant and 2 individuals with the p.(Arg626*) variant. The 6 individuals with SETBP1-RD had missense variants and 4 of which shared the same variant, p.(Glu858Lys) (Table 1). Of note, 12 of the individuals with SETBP1-HD and all 6 of the individuals with SETBP1-RD are not included in prior literature (Figure 1).

**Figure 1:**
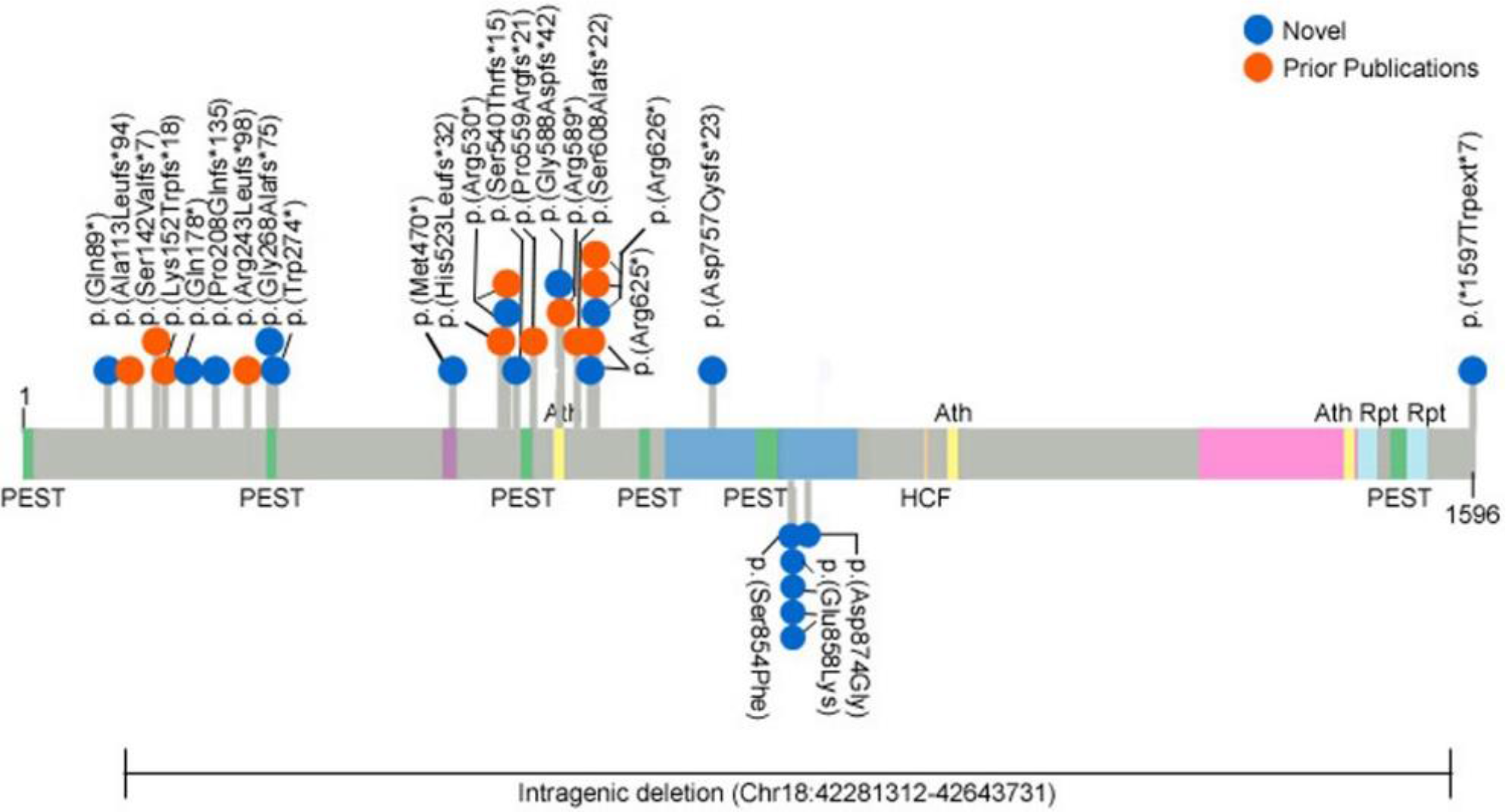
Schematic representation of SETBP1 protein (UniProt: Q9Y6X0), including its domains, indicating SETBP1- HD variants above and SETBP1-RD variants below with blue dots representing novel individuals and orange dots indicating individuals in prior publications. Two individuals with larger deletions (p21, p22) are not shown here. For cDNA annotations of the variants see Table 1.

Six of the 28 SETBP1-HD participants and 1 of the 6 SETBP1-RD participants had no medical history, background, or behavioral results and were not included further in this publication.

### Clinical Results/Clinical Findings

#### SETBP1-HD

Summary clinical characteristics for 22 (15 male and 7 female, age range = 2.2-24.25 years) SETBP1-HD participants are provided (Table 2). Of note, not all individuals completed the MHI or the background history form resulting in missing data (Supplementary Table 2). Speech and language difficulties were reported by 100% of participating individuals based on speech delays, speech disorders, language disorders, or delayed first words (>12 months).^25^ Neurological manifestations were reported including 95% ID/DD, 74% hypotonia, and 26% clumsiness/incoordination. Attention-Deficit/Hyperactivity Disorder (ADHD) and/or medications for attention issues were reported in 65%.^26^ An ASD diagnosis was reported by 21% >= 2 years, with 75% having comorbid ASD and ADHD diagnoses; 0% reported autistic regression.^27^ Autistic traits were noted in 56%; based on elevated scores for one or more of the following: T >= 60 SRS-2 total, T >= 70 on CBCL/6-18 social, attention and thought problems scales, or T >= 65 for the CBCL/1½−5 DSM ASD domain.^20,28^ Sleep issues were reported in 50% and neonatal feeding difficulties in 42%.

**Table 2.**
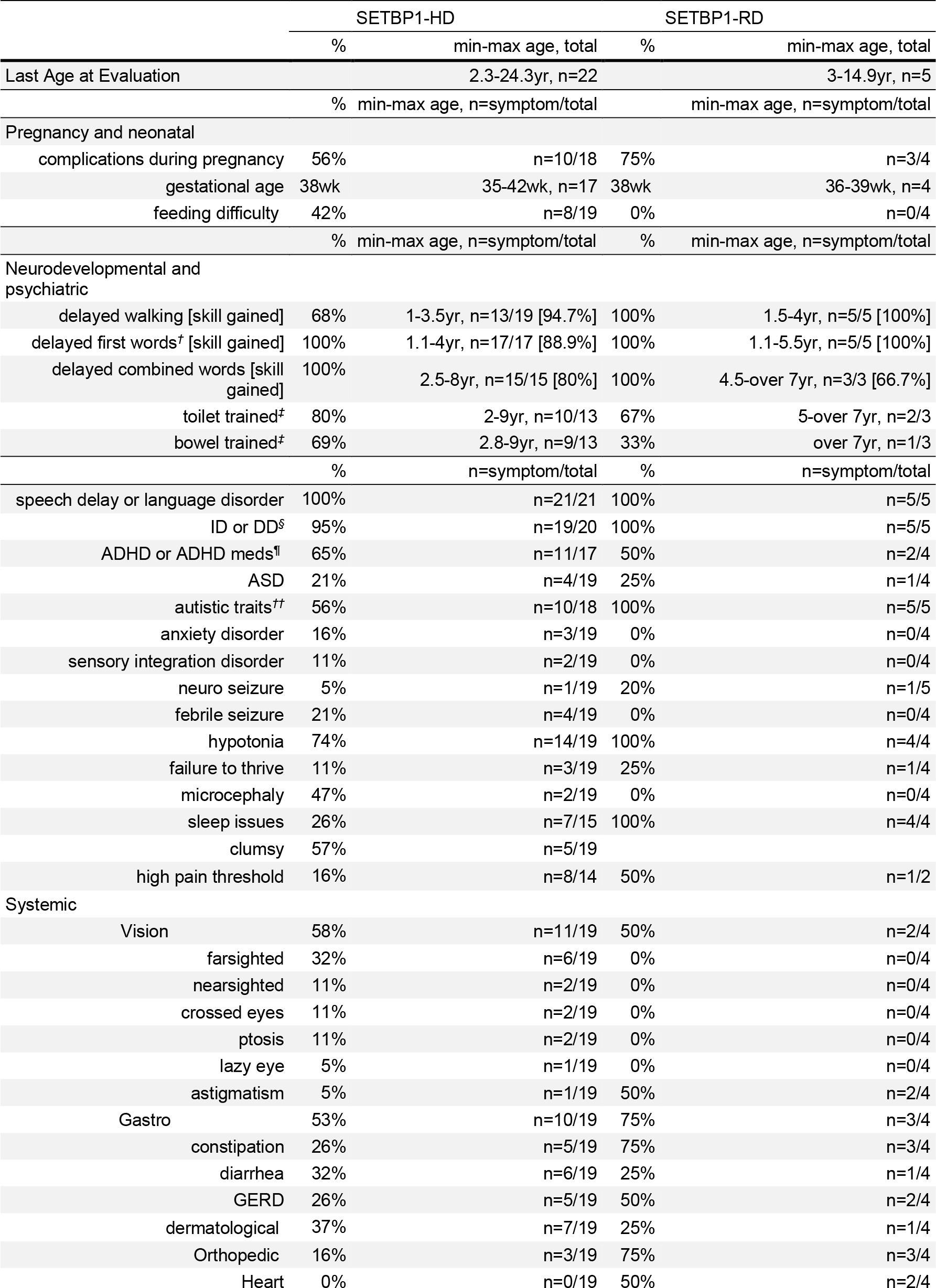
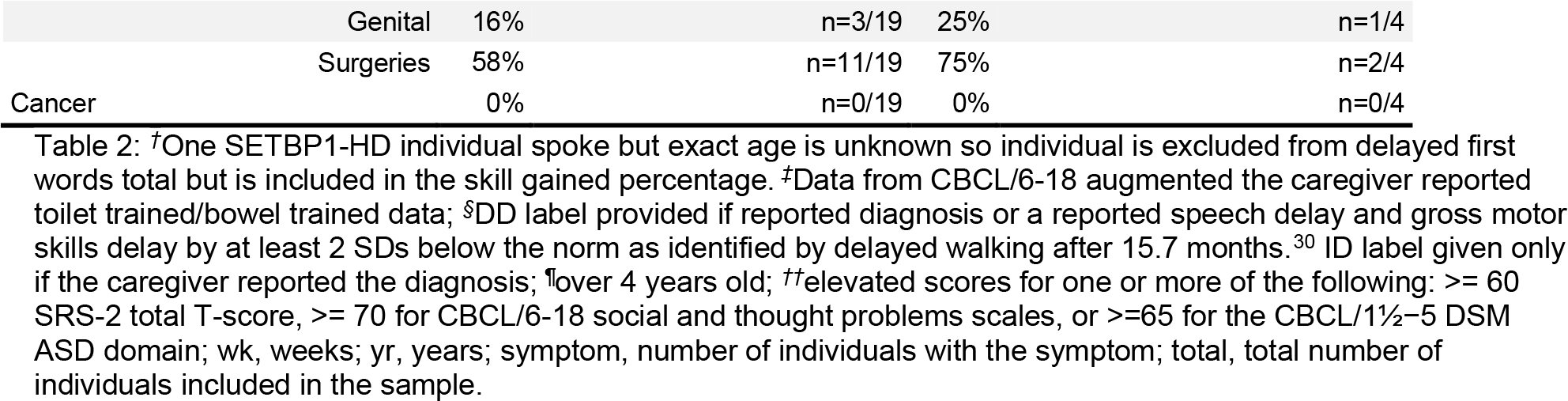
Clinical Characteristics.

In terms of medical issues, 21% of participants reported febrile seizures, of which 1 individual reported an epilepsy diagnosis. A high pain tolerance was reported in 57%, gastrointestinal issues in 53%, failure to thrive in 16%, and anxiety also in 16%. Vision issues were reported in 58%, with farsightedness as the most prevalent issue. Surgeries were noted in 58% with a majority being minor surgeries, including tongue-tie in 21%, PE tubes for 21%, and an adenoidectomy in 14%. Dermatological issues were reported for 37% with eczema being the most commonly reported. Various orthopedic issues were reported by 16%, as well as 16% reporting undescended testicles as the only noted genital issue. Toilet training occurred for 77% with success ranging from 2-9 years of age and bowel control occurred for 68% with success ranging from 2.8-9 years of age.

#### SETBP1-RD

Summary clinical characteristics for 5 (4 female and 1 male, age range = 3-14.9 years) SETBP1-RD participants are provided (Table 2). IDD/DD and speech and language issues as well as hypotonia and sleep issues were identified in 100% of participants. Distinct orthopedic issues were reported in 3 individuals, as well as gastrointestinal issues, with constipation being the most common symptom. Heart issues were reported in 2 individuals. Two individuals reported astigmatism as the only vision issue. Seizures in the form of infantile spasms were reported in 1 individual, with 1 other individual reporting suspected absence seizures. Distinct surgeries were noted in 3 individuals and movement disorders were noted in 2 individuals (Supplementary Table 2). A genital issue of hypospadias was noted in 1 individual. Toilet training occurred for 2 individuals with success ranging from 5 to 7+ years of age and bowel control occurred for 1 individual with success reported at 7+ years of age.

No individuals with SETBP1-HD or SETBP1-RD reported a diagnosis of cancer, which is worth mentioning as somatic SETBP1 variants have been linked to certain blood cancers.^3^ Additionally, individuals with SGS have a higher risk for tumors and malignancy further delineating from the individuals with SETBP1-HD and SETBP1-RD.^6,28^

### Behavioral Results

#### Vineland-3

Vineland-3 data were available for 13 (9 male, age range = 2.3-24 years) SETBP1-HD participants. Borderline to mild impairments were noted across all domains, as reflected by their standard score means. Mild impairment was noted at the group level in the communication (66.7) domain. Borderline impairments were noted for the group in the following domains: socialization (79.3), motor skills (72.4), daily living skills (74.5) and ABC (72.5, range = 59-84). For the subdomain v-scores, expressive (9.7), written (7.2), community (8.7) and fine motor (9.7) fell in the low range for group means (Table 3). At the individual level, scores fell in the moderately low to low range in 82% of the subdomain scores, within the borderline to mild range in 81% of the domain scores, and below the adequate level in 90% of the domain scores.

**Table 3.** SETBP1-HD Vineland-3.

A mean of 100 and SD=15 apply to composite and domain scores, while a mean of 15 and SD=3 apply to subdomain scores.
| VABS-3 Domains (n=13) | Mean (SD) | Median | Range | Impairment Levels |
| --- | --- | --- | --- | --- |
| Communication<sup>†</sup> | 66.7 (12.3) | 73 | 45-83 | Mild |
| Expressive<sup>‡</sup> | 9.7 (3.4) | 10 | 1-15 | Low |
| Receptive<sup>‡</sup> | 10.2 (1.8) | 10 | 6-12 | Moderately Low |
| Written<sup>‡,¶</sup> | 7.2 (3.1) | 8 | 1-10 | Low |
| Daily Living Skills<sup>†</sup> | 74.5 (16.7) | 73 | 54-118 | Borderline |
| Personal<sup>‡</sup> | 10.8 (3.8) | 10 | 7-21 | Moderately Low |
| Domestic<sup>‡,¶</sup> | 11.4 (3.7) | 11 | 7-21 | Moderately Low |
| Community<sup>‡,¶</sup> | 8.7 (2.6) | 9 | 3-12 | Low |
| Social<sup>†</sup> | 79.3 (13.7) | 76 | 56-108 | Borderline |
| Interpersonal Relationships<sup>‡</sup> | 11.5 (2.5) | 11 | 8-15 | Moderately Low |
| Play & Leisure<sup>‡</sup> | 11.7 (3) | 11 | 8-18 | Moderately Low |
| Coping Skills<sup>‡,††</sup> | 10.7 (2.6) | 11 | 7-16 | Moderately Low |
| Motor<sup>†</sup> | 72.4 (5.5) | 73 | 65-79 | Borderline |
| Gross Motor<sup>‡,§</sup> | 10 (1.6) | 9 | 8-12 | Moderately Low |
| Fine Motor<sup>‡</sup> | 9.7 (2.4) | 10 | 7-13 | Low |
| ABC/Composite<sup>†</sup> | 72.5 (8.6) | 71 | 59-84 | Borderline |

Notably, 100% fell below the adequate level in communication (including receptive and written), motor (including gross motor), community within daily living skills, and overall adaptive functioning. An area of relative strength was interpersonal relationships in the socialization domain with 46% in the adequate level (Supplementary Table 3).

The SETBP1-RD group included Vineland-3 data for only 2 individuals (both female, age range = 2.7-9.2 years) with results falling in the SETBP1-HD groups’ defined ranges for each domain (Table 3).

#### Child Behavior Checklist (CBCL)

CBCL/6-18 data were available for 12 SETBP1-HD (10 male and 2 female, age range = 6.3- 14.3 years ) and 3 SETBP1-RD (2 female and 1 male, age range = 10-16 years) participants. CBCL/1 ½-5 data were available for 8 SETBP1-HD (5 female and 3 male, age range = 2.2-6 years) and 3 SETBP1-RD (all female, age range = 3.1-5.5 years) participants.

First, we considered the broadband scales including total problems, internalizing problems, and externalizing problems, as summarized in Figure 2A. For total problems, the mean score was in the borderline range for SETBP1-HD school-age participants (range = 51-77), SETBP1-RD school-age participants (range = 54-74), and SETBP1-RD younger participants (range = 38-81) (Figure 2A).

**Figure 2:**
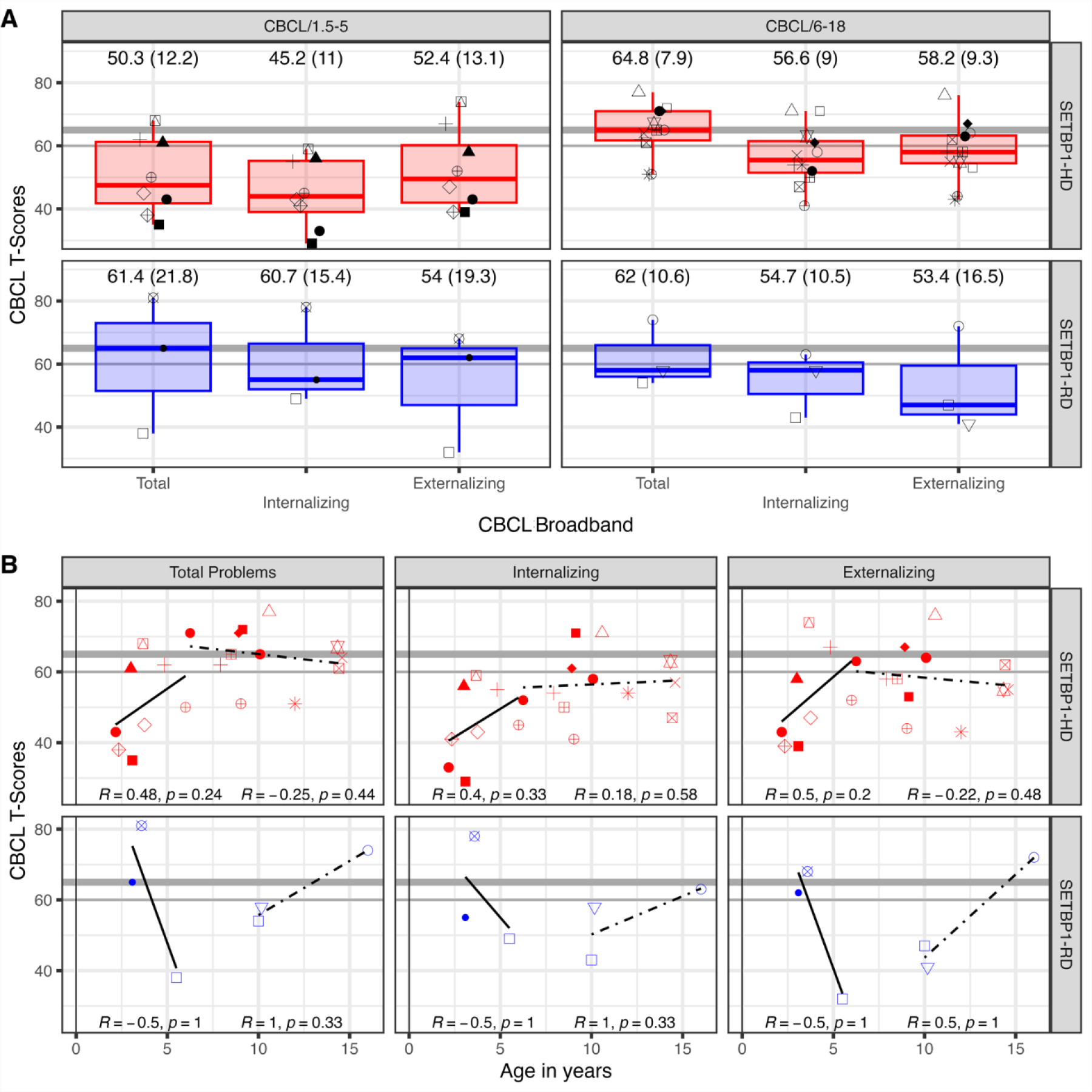
**Cross-sectional effects for broadband scales on Child Behavior Checklists**. The higher the value, the more severe the symptoms. Broadband summary group data provided for internalizing, externalizing, and total with scores of 64+ indicating clinically significant and scores within 60-63 indicating borderline. In Part A, boxplots reflect median and quartile ranges for each group with individual scores represented by the small shapes. The mean and standard deviation scores are represented above the box plots. In Part B: The R and p values represent Spearman correlations between the mean T-score and the individual’s age. The bold lines represent the trend for results on the CBCL/1.5-5 and the dotted lines represent the trend for results on the CBCL/6-18.

A series of ANOVAs of broadband scales evaluated full-factorial effects between SETBP1 group, age, and CBCL measure. An overall age effect, *F*(1, 18) = 5.61, *p* = .030, suggested that total problems increased as children got older; however, an interaction between group, measure, and age, *F*(1, 18) = 10.22, p = .005, indicated that this effect was stronger for SETBP1-HD relative to SETBP1-RD, *p* = .049. A similar pattern was observed for externalizing, *F*(1, 18) = 11.42, *p* = .003. Age effects are illustrated in Figure 2B.

Narrowband scales are described separately in the following sections for each version of the CBCL using similar statistical approaches but without the effect between measurements since models were run separately for CBCL/1.5-5 and CBCL/6-18.

#### CBCL/6-18

For SETBP1-HD, the only clinically significant narrowband scale was attention problems (range = 51-88). Borderline narrowband scales included social problems (range = 58–75), thought problems (range = 59-83) and DSM-oriented scales included DSM ADHD (range = 60-70) (Figure 3).

**Figure 3:**
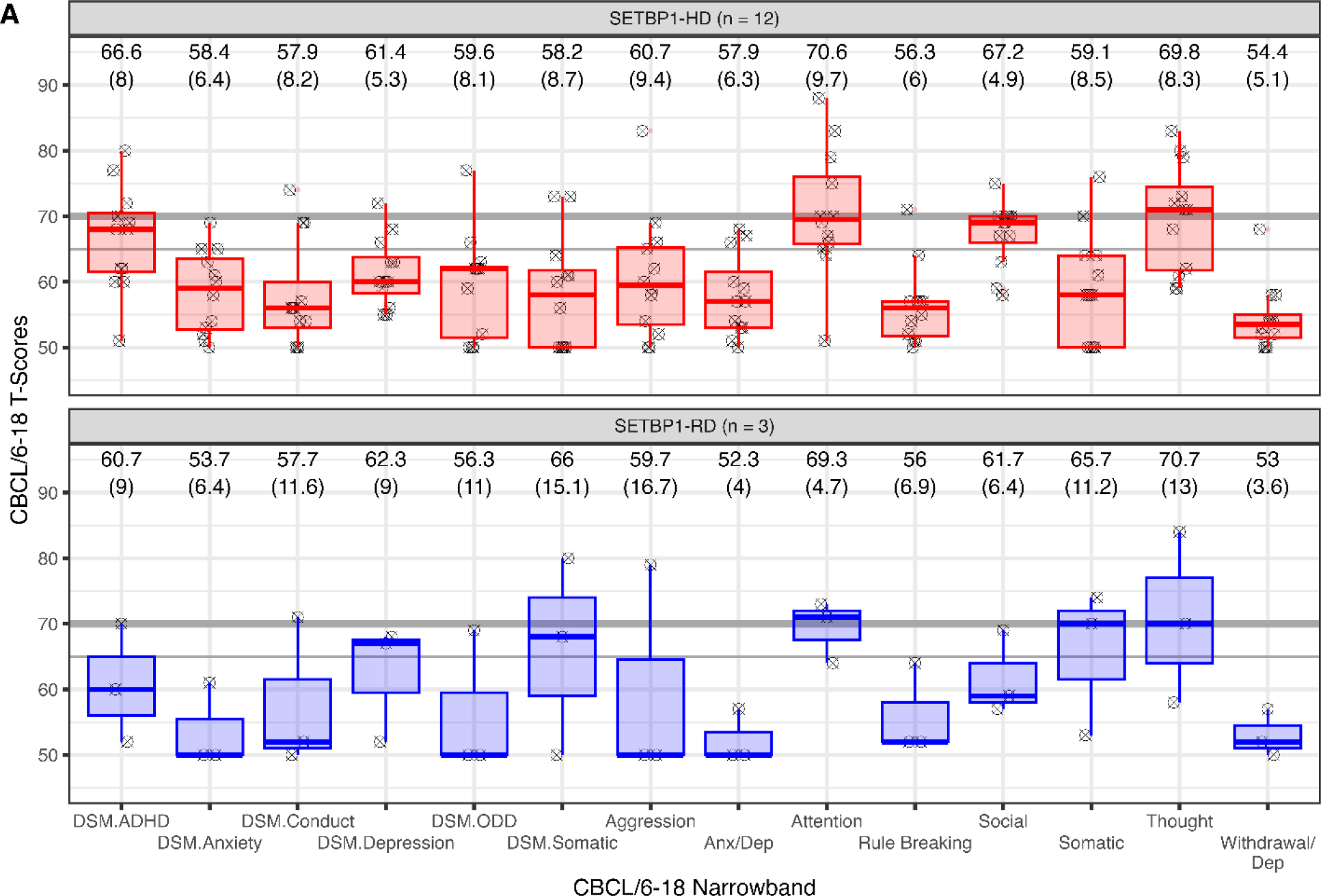
**Cross-sectional effects for narrowband and DSM-oriented scales on Child Behavior Checklist 6-18**. The higher the value, the more severe the symptoms. Summary group data provided for narrowband and DSM- oriented scales with scores of 70+ indicating clinically significant and scores within 65-69 indicating borderline. Boxplots reflect median and quartile ranges for each group with individual scores represented by the small shapes. The mean and standard deviation scores are represented above the box plots.

At the SETBP1-HD individual level, the most common issues reported by 92-100% were easily distracted, speech problems, obsessions, acts young, impulsiveness, clumsy, demands attention, concentration issues, prefers younger kids, picks skin and is too dependent.

For SETBP1-RD results were limited by a small number (*n* = 3). The only clinically significant narrowband scale included thought problems (range = 58-84). Borderline narrowband scales included somatic problems (range = 53-74) and attention problems (range = 64-7) and DSM-oriented scales included DSM somatic problems (range = 50-80).

At the SETBP1-RD individual level, issues reported by 100% of the sample included speech problems, repeats certain acts over and over, acts young, clumsy, concentration issues, bowel movement outside toilet, restless, constipated, and trouble sleeping (Supplementary Table 4).

Main group effects were not observed at the narrowband scale level, p > .11. Group and age interactions were observed for aggressive problems, F(1, 11) = 5.56, p = .038, such that problems increased for SETBP1-RD but not SETBP1-HD participants with age. Similar trends were observed for conduct, rule-breaking, and thought subdomains, p < .079.

#### CBCL/1 ½-5

For SETBP1-HD, the only elevated narrowband scale on the CBCL/1 ½-5 was attention problems (range = 50-80) (Supplementary Table 5). Elevated scores were notable at the individual level including speech problems by 100% and clumsy and gets into everything by 88%.

For the SETBP1-RD cohort, attention problems, withdrawn, and DSM ADHD were all noted at the borderline level for group mean T-scores and with 67% elevated individual scores. With CBCL/1 ½-5 data provided for only 3 individuals with SETBP1-RD, large standard deviations played a significant impact for mean T-scores at the cohort level. At the individual level, speech problems, quickly shifts between activities, acts too young, and resistance to toilet training were indicated by 100%.

Scores were noted as higher for SETBP1-HD relative to SETBP1-RD for DSM anxiety disorders, *F*(1, 7) = 6.79, *p* = .035, and anxiety and depression, *F*(1, 7) = 7.46, *p* = .029.

Interactions between age and SETBP1 group were noted for DSM ADHD, *F*(1, 7) = 8.08, *p* = .025, such that scores decrease more for older SETBP-RD children relative to SETBP1-HD children.

#### SRS-2

Caregivers completed the SRS-2 survey at multiple time points for 5 of the 9 SETBP1-HD participants (18 total surveys completed) and for 2 of the 4 SETBP1-RD participants (10 total surveys completed). For SETBP1-HD participants with SRS-2 data 8 of the 9 were male with an age range of 4.3-14.6 years. The cohort showed moderate elevations for the restrictive and repetitive behaviors (RRB, T M = 72.6), social awareness (SA, T M = 69.4), social cognition (SCog, T M = 72.1), social communication (SCom, T M = 68.5), social communication and interaction (SCI, T M=68.3) and total (T M = 69.9) (Figure 4A). At an individual level, mild to severe social difficulties were identified at 88.9% for RRB, SC, and SCog, at 77.8% for SA, SCI and total, and at a low 33.3% for social motivation.

**Figure 4:**
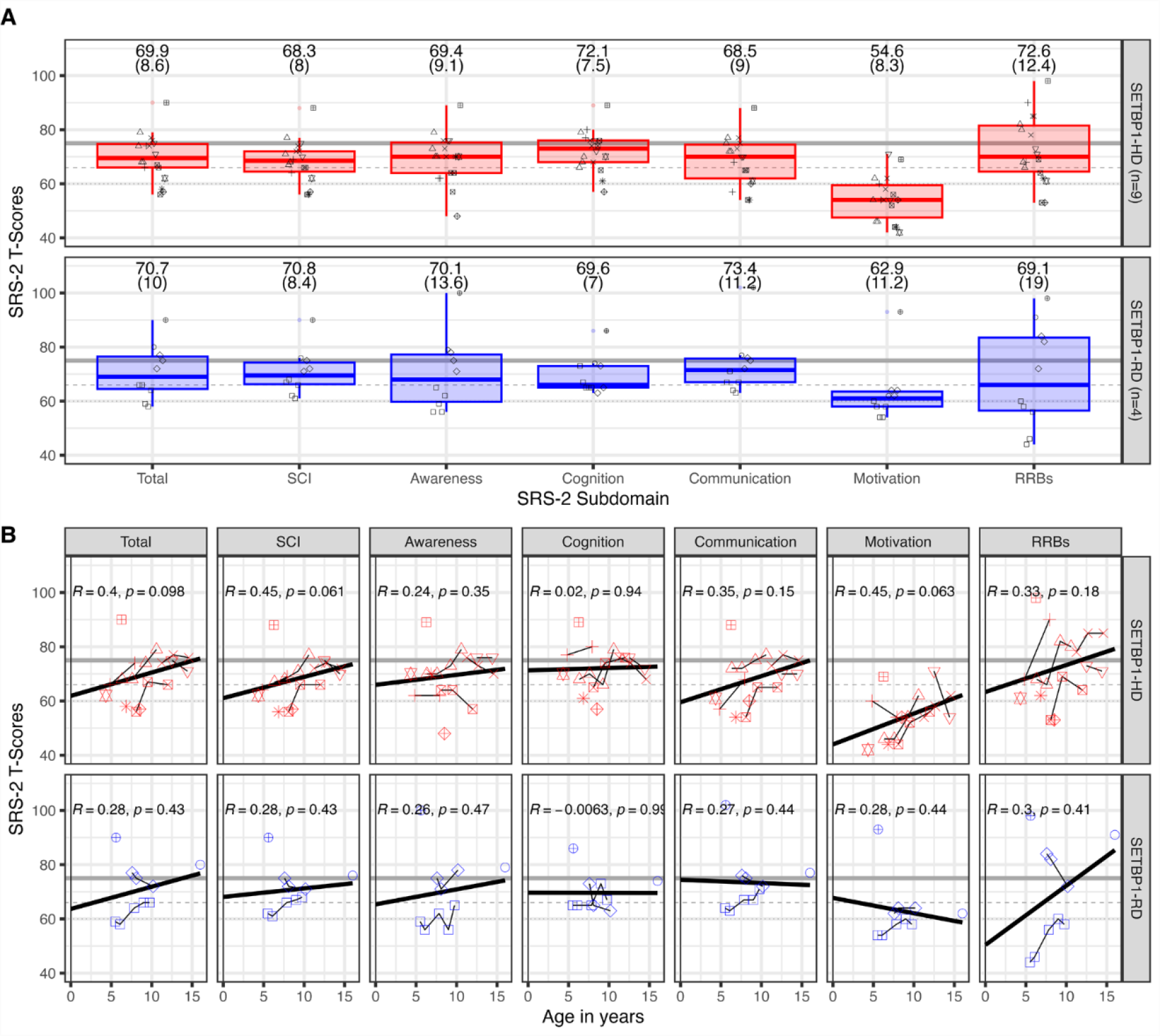
**Cross-sectional and longitudinal effects on SRS-2**. Part A reflects mean T-scores for all Social Responsiveness Scale, Revised (SRS-2) surveys for all subdomains for SETBP1-HD and SETBP1-RD cohorts, respectively. The mean t-scores are stated above the reported standard deviations in parentheses. Scores below the line at 60 fell into the normal range, while scores between the 60 mark and the dotted line at 66 were categorized into mild-moderate range, scores between the dotted line at 66 and the bold line at 75 were categorized into the moderate range and sores above the bold line were categorized into the severe range. Part B plotted all T-scores for each participant representing a mixed cross-sectional/longitudinal graph and connected the scores at each given time point for each participant who completed the SRS-2 at more than one time point to reflect observations over time within SETBP1-HD and SETBP1-RD cohorts, respectively, as shown in above and below charts. The R and p values represented Spearman correlations between the mean T-score and the individual’s age. Note the marginal effect for social communication interaction (SCI) and social motivation for the SETBP1-HD cohort.

For SETBP1-RD participants with SRS-2 data 3 of the 4 were female with an age range of 5.5- 16 years. The cohort reflected scores in the moderate range for the subdomains RRB (T M = 69.1), social awareness (T M = 70.1), social communication (T M = 73.4), social cognition (T M = 69.6), and total SRS-2 (T M = 70.7), and a mild elevation for social motivation (T M = 62.9). Total scores for SRS-2 (Figure 4B) for both SETBP1-HD and SETBP1-RD cohorts were elevated and reflected the overall social deficits observed in symptoms related to ASD.

To compute group differences over time, multilevel linear mixed models with restricted maximum likelihood included a random intercept to account for multiple reports. Longitudinal analysis revealed significant within and between-subject variability for both cohorts. RRB is the only subdomain with a notable increase overtime for the SETBP1-RD cohort (Levene’s test: p = .04). No effects varied by sex. SETBP1-HD and SETBP1-RD cohort differences were marginal for social motivation with scores lower for the SETBP1-HD cohort than the SETBP1-RD cohort: F(1, 24) = 5.046, p = .0342.

#### SCQ

SCQ data were available for 12 SETBP1-HD participants (10 male, age range = 4.8-14.3 years). Based on the responses, 4 were considered nonverbal. Only 1 of the 12 individuals scored above the cutoff for likely ASD >= 15.^21^ This individual has an ASD diagnosis; however, the 2 other individuals with an ASD diagnosis who also completed this survey did not score above the threshold. Lower cutoffs have been suggested in literature for differentiating ASD including a cutoff of >= 10 or 11.^30,31,32^ Six individuals (50%) scored 11 or higher including one individual considered nonverbal, although 1 individual with an ASD diagnosis was still not captured with this cutoff.

SCQ data were available for 3 SETBP1-RD participants (2 female, age range = 5.5-16 years). Based on the responses, all were considered non-verbal. Two of the 3 individuals scored above the cutoff >= 15. One of the individuals with a score above the cutoff had an official ASD diagnosis. All 3 individuals scored above the >= 11 cutoff.

#### CSHQ

CSHQ data were available for 14 SETBP1-HD participants (11 male, age range = 2.2-14.3 years ). With a total sleep disturbance cut-off score of 41, 9 individuals (64.2%) met the criteria to be evaluated for a potential sleep disorder. Additionally, 3 subscales had scores more than 1 standard deviation above the control mean utilized by the original author: night wakings (4.7), parasomnias (10.5), and daytime sleepiness (12.8) (Supplementary Table 6).^22^

CSHQ data were available for 3 SETBP1-RD participants (all female, age range = 5-10.7 years). With a total sleep disturbance cut-off score of 41, 2 individuals (66.6%) met the criteria to be evaluated for a potential sleep disorder. Additionally, 2 subscales were out of range by more than 2 standard deviations above the control mean utilized by the original author; sleep duration (6.6) and night wakings (5.3) and the bedtime resistance (9.3) subscale was out of range by more than 1 standard deviation above the control mean (Supplementary Table 6).

### Medications

During the MHI, medication data on 18 SETBP1-HD participants revealed 61.1% using stimulants and non-stimulants (e.g., methylphenidate, guanfacine) for ADHD symptoms. Gastrointestinal medications were reported by 44%, melatonin for sleep by 16.7%, behavior medications by 16.7%, anti-anxiety meds by 11.1%, and seizure-related meds by 5.6%. Fish oil, excluding multivitamins, was the most common vitamin supplement at 22.2%, with 75% reporting improvement.

Current medication data was collected for 4 SETBP1-RD participants, with 50% reported taking a stimulant or non-stimulant. Seizure-related medication was reported at 50%; however, only 1 individual reported taking them for seizures. Gastrointestinal medications were reported by 75% and 50% reported taking medications for sleep. Fish oil, excluding multivitamins, was the most common vitamin supplement at 50%, with 100% reporting improvement.

## DISCUSSION

Within this paper, we presented the first published findings comparing the neurodevelopmental profiles for individuals with SETBP1-HD and SETBP1-RD. We documented genetic data for 28 individuals with SETBP1-HD and clinical and neurobehavioral details for 22 (12 unpublished).

Additionally, we provided genetic data for 6 individuals with SETBP1-RD and clinical and neurobehavioral details for 5, all previously unpublished cases.

We identified consistent characterization with the SETBP1-HD cohort and previous findings including IDD/DD, speech and language impairment, hypotonia, attention issues, gastrointestinal issues, vision issues, sleep issues, and autistic traits.^1,4,7,14^ These characteristics were also observed in the SETBP1-RD cohort. Notably, both cohorts exhibited a new clinical observation - high pain tolerance. The SETBP1-RD cohort showed clinical comorbidity risks for heart and orthopedic issues, as well as difficulty in bowel control and higher risks for somatic issues compared with the SETBP1-HD cohort. Social motivation emerged as a notable strength for both groups, while repetitive and obsessive behaviors surfaced as a shared challenge. The SETBP1-HD cohort demonstrated relative strength in interpersonal relationships with weaknesses in communication, motor skills, and functioning safely and independently within the community per the VABS-3.

Our findings indicate 100% of SETBP1-HD and SETBP1-RD participants >= 1 year of age reported delayed first words (>12 months) and phrase speech (>24 months).^25^ Communication was reported as the greatest deficit domain on the VABS-3 for the SETBP1-HD cohort, which aligns with a previous study’s findings, and speech problems were indicated as an area of strong weakness on the CBCLs for both cohorts.^4^ The same study mentioned childhood apraxia of speech (CAS) as the most commonly identified speech diagnosis for children with SETBP1- HD at 80% and that the use of language as reported by phonological errors was additionally impacted. Various publications report that individuals with speech and language difficulties are more likely to have difficulty later in other areas, including attention, socialization, reading, and writing.^33,34,35^ According to ASHA, signs of speech and language delays can be identified as early as birth-3 months.^36^ Early detection of language and speech delays and initiation of speech therapy for individuals report positive outcomes in communication and other areas of development for individuals with language delays, ID and/or ASD.^37,38,39,40^

ID/DD were noted for 95% of the individuals with SETBP1-HD and 100% with SETBP1-RD. As a diagnosis of ID now considers adaptive functioning, we also looked at the overall adaptive functioning score on the VABS-3 and noted that all individuals within this study fell in the borderline-mild range.

Despite only 21%-25% reporting an ASD diagnosis for the SETBP1-HD and SETBP1-RD cohorts, the scores for the standardized measures reflect a higher percentage that may warrant further evaluation for an ASD diagnosis and exhibited autistic behaviors, 56% with SETBP1-HD and 100% with SETBP1-RD. Along these same lines, obsessions were noted in all individuals with SETBP1-HD who completed the CBCL/6-18 and repeats certain acts over and over were noted in all individuals with SETBP1-RD who completed the CBCL/6-18. Another predominant characteristic observed was ADHD/attention problems. In the SETBP1-HD cohort, 100% were identified with attention problems (easily distracted, impulsive, and/or concentration issues) on the CBCL/6-18, and 65% reported an ADHD diagnosis and/or were taking medication for associated symptoms. Additionally, 61.1% over the age of 4 reported using stimulants or non-stimulants including methylphenidate and guanfacine. In the SETBP1-RD cohort, 100% were identified with attention problems on the CBCL/6-18, and 50% reported an ADHD diagnosis and/or were taking medication for associated symptoms.

Sleep issues were reported for 47% with SETBP1-HD and 100% with SETBP1-RD. Clinically meaningful differences in sleep disturbances were noted for 64% of individuals with SETBP1- HD in the CSHQ survey and for 2 individuals (67%) with SETBP1-RD. The use of melatonin was reported in both cohorts to aid in sleep problems.

Gastrointestinal issues affected 57% (diarrhea, constipation, and GERD) with SETBP1-HD and 75% (constipation most prevalent) with SETBP1-RD. The elevated Somatic Complaints and DSM Somatic Problems on the CBCL/6-18 for the SETBP1-RD cohort were due to constipation, aches/pains, nausea, stomachaches, and occasional vomiting reported in 67% of individuals.

The SETBP1-RD cohort’s struggle with bowel control as noted in the clinical characteristics and the CBCL/6-18 measure could be linked to the high reports of constipation.

Additional studies are needed to explore how the characteristics of individuals with SETBP1-HD and SETBP1-RD change with age. Additionally, there is a need for studies that specifically isolate and examine the unique neurobehavioral and intelligence differences within each cohort. Although facial differences have been identified for individuals with SETBP1 neurodevelopmental disorders, this data is not captured in Searchlight to include for further phenotyping.^7^ Including the individuals reported in this paper, the number of unique individuals with SETBP1-HD reported in publications increases to 76 (Supplementary Table 7). Presenting the characteristic and phenotype cross-sectional data in this publication for individuals with SETBP1-HD and SETBP1-RD, although limited by a small sample size, is an important step forward for medical management, accurate diagnosis, and clinical trial readiness.

## Data Availability

Approved researchers can obtain the Simons Searchlight population dataset described in this study (https://www.sfari.org/resource/simons-searchlight/) by applying at https://base.sfari.org.

