## Supplemental Table 1 for "SETBP1 Haploinsufficiency and Related Disorders Clinical and Neurobehavioral Phenotype Study"

| **Supplementary Table 1: Individuals with SETBP1 variants in Searchlight** | | | |
| --- | --- | --- | --- |
| Group | Protein Change | Coding Change | Type of Variant |
| SETBP1-HD | p.(Gln89*) | c.265C>T | Nonsense |
| SETBP1-HD | p.(Lys152Trpfs*18) | c.453_454insTGGG | Frameshift |
| SETBP1-HD | p.(Gln178*) | c.532C>T | Nonsense |
| SETBP1-HD | p.(Arg243Leufs*98) | c.726_732del | Frameshift |
| SETBP1-HD | p.(Trp274*) | c.821G>A | Nonsense |
| SETBP1-HD | p.(Met470*) | c.1408del | Nonsense |
| SETBP1-HD | p.(His523Leufs*32) | c.1568del | Frameshift |
| SETBP1-HD | p.(Arg530*) | c.1588C>T | Nonsense |
| SETBP1-HD | p.(Arg530*) | c.1588C>T | Nonsense |
| SETBP1-HD | p.(Ser540Thrfs*15) | c.1619del | Frameshift |
| SETBP1-HD | p.(Pro559Argfs*21) | c.1676del | Frameshift |
| SETBP1-HD | p.(Gly588Aspfs*42) | c.1763del | Frameshift |
| SETBP1-HD | p.(Arg589*) | c.1765C>T | Nonsense |
| SETBP1-HD | p.(Ser608Alafs*22) | c.1821del | Frameshift |
| SETBP1-HD | p.(Arg625*) | c.1873C>T | Nonsense |
| SETBP1-HD | p.(Arg625*) | c.1873C>T | Nonsense |
| SETBP1-HD | p.(Arg625*) | c.1873C>T | Nonsense |
| SETBP1-HD | p.(Arg626*) | c.1876C>T | Nonsense |
| SETBP1-HD | p.(Arg626*) | c.1876C>T | Nonsense |
| SETBP1-HD | p.(Asp757Cysfs*23) | c.2269_2281del | Frameshift |
| SETBP1-HD | SETBP1 Deletion | arr[GCRh37] 18q12.3 (40846809-42883959)x1 | Deletion |
| SETBP1-HD | SETBP1 Deletion | arr[GRCh37] 18q12.3 (40034922-42755922)x1 | Deletion |
| SETBP1-HD | p.(Ala113Leufs*94) | c.337del | Frameshift |
| SETBP1-HD | p.(Ser142Valfs*7) | c.422 dup | Frameshift |
| SETBP1-HD | p.(Pro208Glnfs*135) | c.623del | Frameshift |
| SETBP1-HD | p.(Gly268Alafs*75) | c.801del | Frameshift |
| SETBP1-HD | p.(*1597Trpext*7) | c.4790_*9del | Frameshift |
| SETBP1-HD | Deletion of exon 2-6 | arr[GCRh37] 18q12.3 (42281312-42643731)x1 | Deletion |
| SETBP1-RD | p.(Ser854Phe) | c.2561C>T | Missense |
| SETBP1-RD | p.(Glu858Lys) | c.2572G>A | Missense |
| SETBP1-RD | p.(Glu858Lys) | c.2572G>A | Missense |
| SETBP1-RD | p.(Glu858Lys) | c.2572G>A | Missense |
| SETBP1-RD | p.(Asp874Gly) | c.2621A>G | Missense |
| SETBP1-RD | p.(Glu858Lys) | c.2572G>A | Missense |
| VUS | p.(Arg67Trp) | c.199C>T | VUS (unknown) |
| VUS | p.(Arg67Trp) | c.199C>T | VUS (inherited) |
| VUS | p.(Gly64Asp) | c.191G>A | VUS (unknown) |
| VUS | p.(Lys435Glu) | c.1303A>G | VUS (unknown) |
| VUS | p.(Lys469Gln) | c.1405A>C | VUS (unknown) |
| VUS | p.(Lys469Gln) | c.1405A>C | VUS (inherited) |
| VUS | p.(Ser772Leu) | c.2135C>T | VUS (unknown) |
| VUS | p.(Ser772Leu) | c.2135C>T | VUS (unknown) |
| SGS | p.(Gly870Arg) | c.2608G>C | SGS (unknown) |
| SGS | p.(Gly870Arg) | c.2608G>C | SGS (inherited) |
| SGS | p.(Ile871Ser) | c.2612 T>G | SGS (de novo) |
| VUS | p.(Asp900Gly) | c.2699A>G | VUS (inherited) |
| VUS | p. (Leu957Pro) | c.2870T>C | VUS (de novo) |
| VUS | p.(His1167Asn) | c.3499C>A | VUS (unknown) |
| VUS | p.(Arg1146Trp) | c.3436C>T | VUS (inherited) |
| VUS | p.(Arg1146Trp) | c.3436C>T | VUS (unknown) |
| VUS | p.(His1158Leu) | c.3473A>T | VUS (unknown) |
| VUS | p.(Trp1242Arg) | c.2734T>C | VUS (unknown) |
| VUS | p.(Cys1478Gly) | c.4432T>G | VUS (unknown) |
| VUS | p.(Cys1478Gly) | c.4432T>G | VUS (inherited) |
| VUS |  | arr[GCRh37] 18q12.3 (42519449-42567640)x1 | VUS (unknown) |
| Large Deletion |  | arr[GRCh37] 18q12.3q21.1 (39836056_44849232)x1 | Large deletion including numerous genes |
| Large Deletion |  | arr[GRCh37] 18q12.3q21.1 (39836056_44849232)x1 | Large deletion including numerous genes |
| Large Deletion |  | arr[GRCh37] 18q12.2-12.3 (33777232-42365242) | Large deletion including numerous genes |
| Splice Site |  | c.4000+2T>G | Splice site, no data |
