## Supplemental Table 3 for "SETBP1 Haploinsufficiency and Related Disorders Clinical and Neurobehavioral Phenotype Study"

| **Supplementary Table 3: Individual Vineland-3 Data for SETBP1-HD and SETBP1-RD participant** | | | | | | | | | | |  |  | |  | |  | |
| --- | --- | --- | --- | --- | --- | --- | --- | --- | --- | --- | --- | --- | --- | --- | --- | --- | --- |
| No | sex | age-yy:mm | communication_standard | communication_adaptive_level | motor_standard | motor_adaptive_level | dls_standard | | dls_adaptive_level | soc_standard | | | soc_adaptive_level | | abc_standard | | abc_adaptive_level |
| p2 | male | 0-5yrs | 83 | Borderline | 73 | Borderline | 69 | | Mild | 108 | | | Adequate | | 83 | | Borderline |
| p4 | male | 6-10yrs | 74 | Borderline | 76 | Borderline | 70 | | Borderline | 72 | | | Borderline | | 71 | | Borderline |
| p6 | male | 6-10yrs | 76 | Borderline | 76 | Borderline | 76 | | Borderline | 94 | | | Adequate | | 79 | | Borderline |
| p7 | male | 6-10yrs | 73 | Borderline | - |  | 65 | | Mild | 68 | | | Mild | | 68 | | Mild |
| p9 | female | 6-10yrs | 73 | Borderline | - |  | 81 | | Borderline | 85 | | | Borderline | | 77 | | Borderline |
| p10 | female | 21-25yrs | 57 | Mild | - |  | 54 | | Moderate | 56 | | | Mild | | 59 | | Mild |
| p14 | male | 11-15yrs | 49 | Moderate | - |  | 54 | | Moderate | 76 | | | Borderline | | 62 | | Mild |
| p16 | male | 11-15yrs | 79 | Borderline | - |  | 78 | | Borderline | 72 | | | Borderline | | 75 | | Borderline |
| p17 | male | 6-10yrs | 61 | Mild | 65 | Borderline | 118 | | Mildy High | 84 | | | Borderline | | 84 | | Borderline |
| p18 | female | 6-10yrs | 53 | Moderate | 73 | Borderline | 73 | | Borderline | 76 | | | Borderline | | 67 | | Mild |
| p19 | male | 21-25yrs | 68 | Mild | - |  | 73 | | Borderline | 71 | | | Borderline | | 70 | | Borderline |
| p21 | female | 0-5yrs | 76 | Borderline | 79 | Borderline | 92 | | Adequate | 95 | | | Adequate | | 84 | | Borderline |
| p22 | male | 0-5yrs | 45 | Moderate | 65 | Borderline | 65 | | Mild | 74 | | | Borderline | | 63 | | Mild |
| p30 | female | 6-10yrs | 59 | Mild | 71 | Borderline | 57 | | Mild | 64 | | | Mild | | 63 | | Mild |
| p32 | female | 0-5yrs | 67 | Mild | 73 | Borderline | | 75 | Borderline | 73 | | | Borderline | | 71 | | Borderline |
