## Supplemental Table 5 for "SETBP1 Haploinsufficiency and Related Disorders Clinical and Neurobehavioral Phenotype Study"

| **Supplementary Table 5: CBCL/1.5-5 Narrowband** | | | | | | | | | |  |  | |  |  | |  | |  | |
| --- | --- | --- | --- | --- | --- | --- | --- | --- | --- | --- | --- | --- | --- | --- | --- | --- | --- | --- | --- |
| SETBP1-HD (Participants n = 8) | | | | | | | | | | |  | | SETBP1-RD (Participants = 3) | | | | | | |
| IMPAIRMENT LEVELS | SD | RANGE | | | MEDIAN | | MEAN | | CBCL (1.5-5) | | | MEAN | | MEDIAN | RANGE | | SD | | IMPAIRMENT LEVELS |
| Typical | 0.5 | 50-51 | | | 50 | | 50.4 | | Anxiety/Depression | | | 58.7 | | 56 | 50-70 | | 10.3 | | Typical |
| Typical | 6.8 | 50-70 | | | 53.5 | | 54.9 | | Withdrawn | | | 69.3 | | 67 | 56-85 | | 14.6 | | Borderline |
| Typical | 4.2 | 50-62 | | | 50 | | 52.3 | | Somatic Complaints | | | 57.7 | | 53 | 50-70 | | 10.8 | | Typical |
| Typical | 4.8 | 50-62 | | | 50.5 | | 53.4 | | Emotional Problems | | | 59.3 | | 55 | 50-73 | | 12.1 | | Typical |
| Typical | 9.1 | 50-76 | | | 50.5 | | 55.3 | | Sleep Problems | | | 67.3 | | 64 | 50-88 | | 19.2 | | Borderline |
| Borderline | 12.5 | 50-80 | | | 67 | | 65.1 | | Attention Problems | | | 66.7 | | 70 | 50-80 | | 15.3 | | Borderline |
| Typical | 8.1 | 50-69 | | | 50 | | 54.5 | | Aggressive | | | 57.3 | | 59 | 50-63 | | 6.7 | | Typical |
| Typical | 6.2 | 50-63 | | | 55.5 | | 56.3 | | Stress | | | 63.7 | | 63 | 50-78 | | 14.0 | | Typical |
| Typical | 3.4 | 50-60 | | | 51 | | 51.9 | | DSM Depression | | | 62.7 | | 56 | 50-82 | | 17.0 | | Typical |
| Typical | 3.5 | 50-60 | | | 50 | | 51.4 | | DSM Anxiety Disorder | | | 61 | | 60 | 50-73 | | 11.5 | | Typical |
| Typical | 9.0 | 50-72 | | | 52.5 | | 57.1 | | DSM Autism | | | 68.7 | | 64 | 61-81 | | 10.8 | | Borderline |
| Typical | 9.2 | 50-76 | | | 58.5 | | 59.5 | | DSM ADHD | | | 65.7 | | 71 | 50-76 | | 13.8 | | Borderline |
| Typical | 5.5 | 50-64 | | | 50 | | 52.9 | | DSM ODD | | | 53.3 | | 55 | 50-55 | | 2.9 | | Typical |
| CBCL/1.5-5; Child Behavior Checklist 1.5-5, n; number, SD; standard deviation | | | | | | | | | | | | |  |  | |  | |  | |
