## Supplemental Table 6 for "SETBP1 Haploinsufficiency and Related Disorders Clinical and Neurobehavioral Phenotype Study"

| **Supplementary Table 6: CSHQ** | | | |  |  |  | | |  |  | |  |  |  |
| --- | --- | --- | --- | --- | --- | --- | --- | --- | --- | --- | --- | --- | --- | --- |
|  |  | | SETBP1-HD | | | | | | | SETBP1-RD | | | | |
|  |  | | Control Sample | | | Community Sample | | |  | Control Sample | | | Community Sample |  |
| No | Subscales | mean | | | SD | | mean | diff | | mean | SD | | mean | diff |
| 1 | Bedtime Resistance | 7.06 | | | 1.9 | | 7.85 |  | | **7.06** | 1.89 | | 9.25 | >1 SD |
| 2 | Sleep Onset Delay | 1.25 | | | 0.5 | | 1.42 |  | | 1.25 | 0.53 | | 1.25 |  |
| 3 | Sleep Duration | 3.41 | | | 0.9 | | 3.85 |  | | **3.41** | 0.93 | | 5.75 | >2 SD |
| 4 | Sleep Anxiety | 4.89 | | | 1.5 | | 6.00 |  | | 4.89 | 1.45 | | 5.253 |  |
| 5 | Night Wakings | **3.51** | | | 0.9 | | 4.77 | >1 SD | | **3.51** | 0.89 | | 5.57 | >2 SD |
| 6 | Parasomnias | **8.11** | | | 1.3 | | 10.56 | >1 SD | | 8.11 | 1.25 | | 8 |  |
| 7 | Sleep Disordered Breathing | 3.24 | | | 0.6 | | 3.77 |  | | 3.24 | 0.63 | | 3.5 |  |
| 8 | Daytime Sleepiness | **9.64** | | | 2.8 | | 12.85 | >1 SD | | 9.64 | 2.8 | | 11.416 |  |
| diff; difference | | |  | |  |  | | |  |  | |  |  |  |
